# Detection of mesial temporal lobe epilepsy with OP-MEG

**DOI:** 10.1101/2024.10.17.24315226

**Authors:** Stephanie Mellor, George C. O’Neill, Daniel Bush, Arjun Ramaswamy, David Doig, Tim M. Tierney, Meaghan E. Spedden, Matthew C. Walker, Gareth R. Barnes, Umesh Vivekananda

## Abstract

**Background and Objectives:** Magnetoencephalography (MEG) using optically pumped magnetometers (OP-MEG) is a relatively novel neuroimaging modality that holds great clinical promise for presurgical planning in epilepsy. However, there is limited data demonstrating that interictal discharges from deep neural sources can be visually identified and localised using OP-MEG. In this study, we sought to demonstrate the potential of OP-MEG for recording interictal epileptiform activity from patients with mesial temporal lobe epilepsy, the most common focal epilepsy, with a diverse range of aetiologies.

**Methods:** We recorded whole-head OP-MEG for a minimum of 30 minutes from 8 patients with temporal lobe epilepsies. Patients were seated with their head unconstrained. For comparison, we collected information from previous clinical assessment, including findings from MRI and EEG telemetry.

**Results:** We observed interictal epileptiform activity in 4 of the patients, in 3 of which the localisation of the activity was concordant with previous clinical MRI and EEG. In 2 of those patients, we also observed ictal events. Of those 2 patients, one had a clear abnormality in MRI, with which the localisation of this ictal activity was concordant. The other patient’s MRI was negative, but the ictal localisation is consistent with the seizure semiology.

**Discussion:** We demonstrate that interictal and ictal epileptiform activity from mesial temporal lobe epilepsies can be observed with OP-MEG and localised to the region of the anatomical lesion, identified in MRI. This affirms the utility of OP-MEG for epilepsy surgery planning.

## 1 Introduction

Mesial temporal lobe epilepsy (mTLE), often associated with hippocampal sclerosis, is the most common focal epilepsy that does not respond to medication^1^. An alternative treatment approach for drug-resistant mTLE is resective surgery i.e. removal of mesial temporal structures, which results in seizure freedom in up to 75% of cases^2,3^. Electroencephalography (EEG), which can detect abnormal interictal or ictal electrophysiological activity, and Magnetic Resonance Imaging (MRI) are the mainstay investigations for consideration of surgery. The demonstration by MRI of a lesion, obvious atrophy, or altered signal intensity of mesial temporal structures is extremely valuable in the presurgical evaluation of patients with temporal lobe epilepsy (TLE). However, the ability of scalp EEG to detect and accurately source localise mesial temporal epileptiform activity remains controversial^4^.

Magnetoencephalography (MEG) is a non-invasive functional imaging modality which, like EEG, is a direct measure of electrophysiology. Magnetic fields from, predominantly, post-synaptic potentials are measured and localised. The localisation of MEG signals is less dependent on individual anatomy than EEG, and so can potentially be more accurate. Resecting the epileptogenic focus identified with MEG is a strong predictor of long-term seizure freedom post-surgery^5^. However, MEG has traditionally been limited by the cryogen required for the sensors, resulting in a fixed system with a large footprint. The fixed nature of the system means that patient movement is highly restricted, constrained to within a fixed sensor array, limiting participant compliance. MEG using optically pumped magnetometers (OP-MEG) can overcome some of the limitations of traditional MEG^6^, allowing a system with a form-factor and movement tolerance more comparable with scalp EEG. OP-MEG has been shown to increase the observed SNR of interictal epileptiform discharges (IEDs) in school-aged children by comparison with traditional MEG^7^.

OP-MEG of mTLE and deep sources like the hippocampus is, however, potentially more challenging. OPMs can be placed directly on the scalp, closer to the epileptogenic focus than previous MEG sensors and so the signal strength is higher. However, the percentage increase in signal decreases with depth within the brain, and the measurement of deep sources such as the hippocampus remains controversial within traditional MEG literature^8^. Previous data from a single patient suggests that mTLE data can, nevertheless, be recorded with OP-MEG^9^. In this study, we analysed 30-minute OP-MEG recordings from 4 patients with varying temporal lobe pathologies. We demonstrate clear interictal and ictal epileptiform activity, recorded from adults sitting with their head unconstrained.

## 2 Methods

### 2.1 Patient Information

8 adult patients (2 female) participated in this study. No interictal or ictal events were observed in 4 patients (1 female) and so they were excluded from further analysis. Demographics of the remaining 4 patients can be found in Table 1. Ethical approval for the study was given by the Essex Research Ethics Committee in the UK (REC Reference 18/EE/0220), and informed consent was obtained from all patients prior to participation.

**Table 1.**
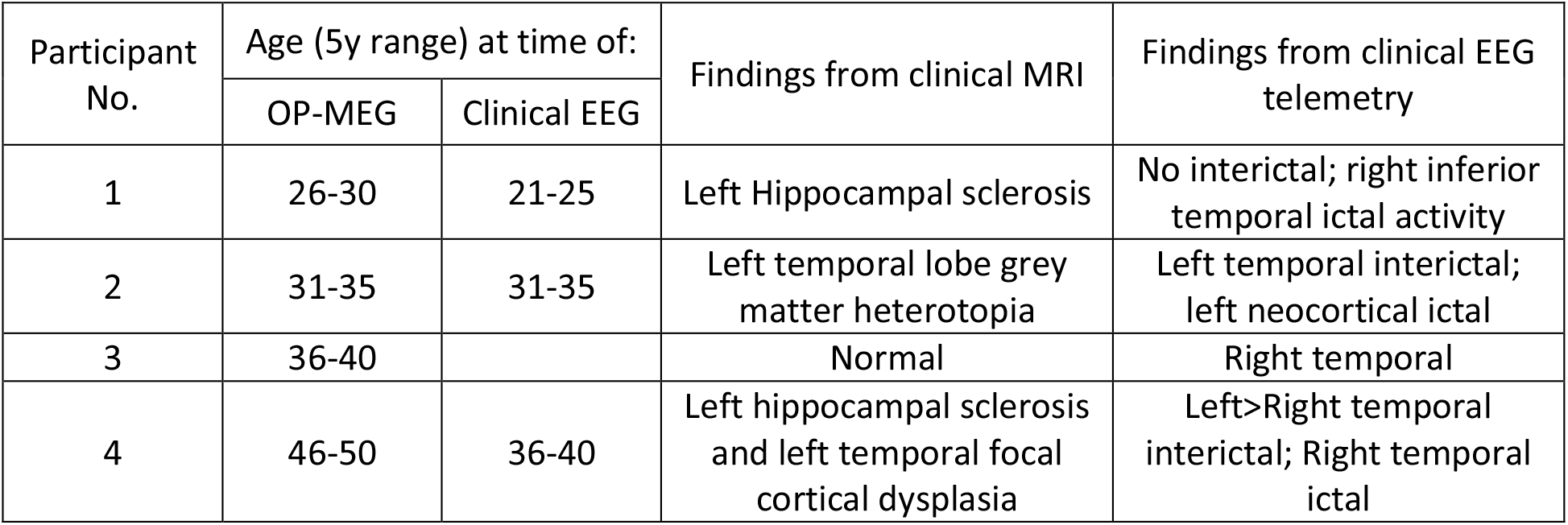
Patient demographics and clinical characteristics.

Patients were not preselected for presence of IEDs in previous electroencephalography (EEG). All patients had routine EEGs within normal limits in their standard clinical assessment, therefore necessitating prolonged recording. The clinical EEG telemetry of the 4 patients rejected from further analysis showed no ictal epileptiform activity or interictal spiking for 2 patients (one of whom did exhibit interictal slow wave activity in the EEG). One patient exhibited temporal/frontotemporal interictal sharp waves and non-lateralising seizure activity, while the remaining patient exhibited right sphenoidal interictal activity and right temporo-parietal region ictal activity.

### 2.2 Data collection

#### 2.2.1 OP-MEG system

All data collection was performed within the OPM Magnetically Shielded Room (MSR) at UCL (Magnetic Shields Limited, 4-layer MSR, internal dimensions 3 m x 4 m x 2.2 m). The number of OPMs and sensor arrangement was different for each patient (see Figure 1) but in all cases, an array of zero-field OPMs (Gen-2.0 and Gen-3.0 QZFM, QuSpin Inc., Louisville, CO) was held on the patient’s scalp in a bespoke 3D-printed scanner-cast, designed from the patient’s MRI ^10^. OPM data were recorded via a 16-bit analogue to digital converter (NI-9205, National Instruments; Austin, TX) at 6 kHz. The Gen-2.0 OPMs were operated in dual-axis mode (meaning the field component radial to the scalp and one axis tangential to it was recorded), while the Gen-3.0 OPMs recorded all three magnetic field components. Here we will use the term sensor or OPM to refer to a single device and channel to refer to the recorded data from one axis of that device, i.e. one OPM had 2 or 3 channels. For patient 1, the dynamic range of each channel was set to ±1.9 nT. Otherwise, the dynamic range of each channel was ±5.56 nT.

**Figure 1.**
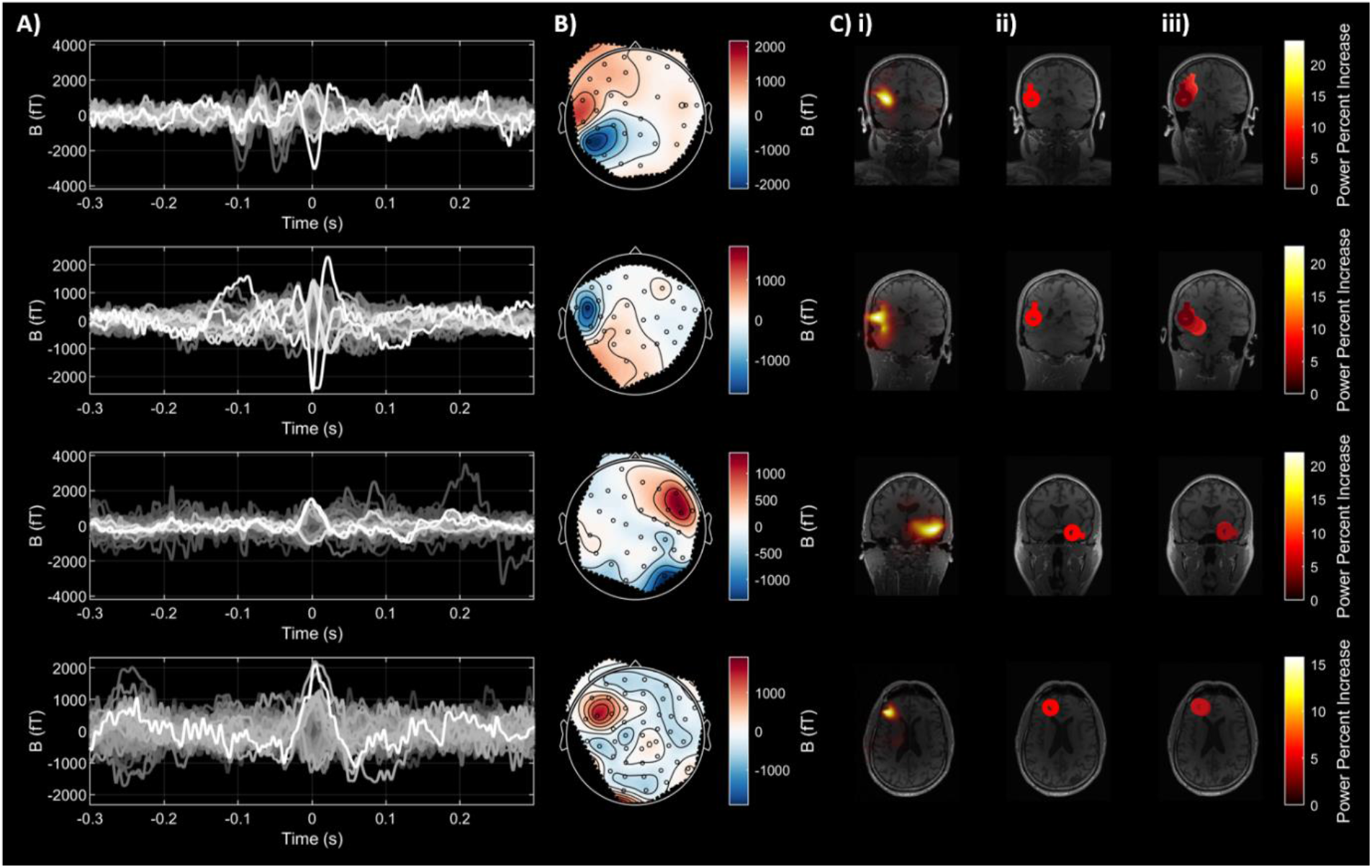
Interictal results from all 4 patients. A) Time series of the average spike. B) Topography of the average spike projected onto a pseudo-10-20 layout. C) Source Localisation results. i) LCMV beamformer, ii) Dipole fit of peak, iii) Dipole fits along the rising edge of the average spike, taken at 2 ms intervals starting -10 ms. The colours get darker going forward in time.

For patients 1-3, only Gen-2.0 OPMs were used (38, 42 and 38 respectively). For patient 4, 32 Gen-2.0 and 25 Gen-3.0 OPMs were used. The variation in the number of sensors was due to the development of the OP-MEG system as the recordings were being undertaken. To minimise the impact of this and allow for better comparison between patients, when plotting sensor-level topographies, sensor positions have been projected into a pseudo-10-20 space (https://github.com/spm/spm/blob/main/spm_get_anatomical_layout.m). Up to 4 OPMs were later rejected from each recording due to excessive noise in the data; full numbers are given in the supplementary information. Two further Gen-2.0 OPMs were removed from the analysis of patient 2’s ictal data, as there was high noise in these channels for this specific time period of interest. Retro-reflective markers were also placed on the scanner-cast to record the patients’ position and rotation from an array of 6 OptiTrack Flex 13 cameras (Natural Point Inc., Corvallis, OR) sampled at 120 Hz.

Prior to recording, the MSR was degaussed ^11^. The patients were asked to sit as still as possible for 30 s while the currents through the on-board OPM coils were optimised to minimise remnant background fields and the OPMs were calibrated following the manufacturer’s procedure. No additional active shielding was used and OPMs were open-loop, i.e. these on-board coil currents were set before the start of the experiment and kept constant throughout the recording.

#### 2.2.2 Paradigms

Each patient sat on a beanbag in the centre of the MSR with their head unconstrained for approximately 1 hour. The OPM cables were fixed to a table behind the patient in order to reduce the weight on the patient’s head but allowing head movement of at least 20 cm in any direction. A clinician was also in the room for recordings from patients 2, 3 and 4.

The patients performed a range of tasks, including a memory, reward processing and language paradigm. Details of each paradigm and which patients undertook it are given in the supplementary material. Additionally, for patients 1, 2 and 4, at least 10 minutes of data (exact timing given in supplementary table 1) were recorded while the patients were not explicitly performing a task. For patients 2 and 4, this was recorded while the participant talked to the clinician. Patient 1 instead read a book. Patient 3 had musicogenic epilepsy, and so a 10 minute recording was instead undertaken while listening to music intended to induce a seizure. We do not present the cognitive data here but do show interictal spikes recorded while the participants undertook the tasks. The number of IEDs observed in each recording separately is given in the supplementary material; the results presented here are averaged over recordings.

### 2.3 Data Analysis

#### 2.3.1 Pre-processing

All analysis was performed in SPM ^12^ (https://github.com/spm/spm) unless otherwise stated. OPM data were first down-sampled to 1 kHz for computational efficiency and trimmed to only the period when the motion tracking was simultaneously recording. The motion tracking data were then upsampled using linear interpolation to match the OP-MEG data at 1 kHz. Based on the power spectral density, anomalous channels were then visually identified and rejected from further analysis.

Six 5^th^ order Butterworth filters were applied bidirectionally to the OPM data. Band-stop filters at 50 Hz, 100 Hz and 150 Hz were included to remove line noise, as well as a band-stop filter at 120 Hz to remove interference from the OptiTrack cameras. The data were then low-pass filtered with a cut-off of 130 Hz and high-pass filtered with a cut-off frequency of 3 Hz. Spatial filtering was then used to further minimise background interference. These separate external interference from the brain signals of interest based on their spatial patterns. If the data contained over 99 good MEG channels, Adaptive Multipole Modelling (AMM) ^13^ was used with the temporal extension with a correlation limit of 0.95. Otherwise, Homogeneous Field Correction (HFC) ^14^, a comparatively simpler model requiring fewer channels, was used. Independent Component Analysis (ICA) was then used to remove heartbeat artefacts and eyeblinks. The number of components removed for each recording is given in the supplementary material.

#### 2.3.2 Spike detection

An experienced clinician (UV) and OP-MEG researcher (SM) then visually examined the preprocessed data and the ICA components for interictal spikes and artefacts.

Initially, no spikes were identified in the sensor-level data for patient 3. To further reduce background interference, a Linearly Constrained Minimum Variance (LCMV) beamformer was then used to create virtual electrodes for each area of the AAL atlas as follows. The beamformer weights for a 5 mm grid within the patient’s inner-skull surface were estimated in SPM using the DAiSS toolbox. The pre-processed OPM data were low-pass filtered at 70 Hz (5^th^ order Butterworth filter applied bidirectionally), then epoched into 5 s trials (to reduce matrix size for covariance computation). The covariance matrix was truncated to 62 components (the number of degrees of freedom after denoising with HFC and ICA). To calculate the lead fields, the Nolte single shell forward model was used ^15^. To estimate the spatial filter for each area of the AAL atlas, we identified the tissue type of each grid point by interpolating the AAL atlas with FieldTrip and then used singular value decomposition to obtain a single spatial filter for each region from the filters for each grid point in the region. UV then visually examined these virtual electrodes to identify 4 IEDs for patient 3. These were seen in multiple AAL parcellations, including the cerebellum, frontal regions and the hippocampal, parahippocampal and insular regions. These time points were then used to epoch the sensor-level OPM data, and source analysis was undertaken as for the other patients.

#### 2.3.3 Source localisation

To evaluate the OP-MEG performance against previous clinical assessment, source localisation results from an LCMV Beamformer and Equivalent Current Dipole Fitting were evaluated from the pre-processed data without ICA applied. In both cases, the Nolte single shell forward model was used ^15^.

##### 2.3.3.1 Beamformer

The LCMV Beamformer was implemented in the same way as in section 2.3.2 with the following changes. For the covariance matrix, for all but patient 2, 40 seconds of data centred on each spike were combined and filtered between 2 Hz and 90 Hz. For patient 2, 8 s of data centred on each spike were used, as more spikes were available and so less data per spike was required for there to be sufficient data overall to estimate the covariance matrix, allowing the inclusion of a spike observed only 4.4 s into a recording. The truncation of the covariance matrix for regularisation varied between recordings due to the varying number of sensors. For patients 1, 2, 3 and 4 respectively, 64, 70, 64 and 90 components were used. A baseline power image was calculated from the time range 1.1 s – 1 s before each spike. A power image was also calculated for the 100 ms centred on each spike, and a contrast calculated as the percentage power change.

##### 2.3.3.2 Dipole Fitting

Dipole fitting was performed in the FieldTrip toolbox. The individual spikes were averaged and dipoles fitted to the peak and rising edge of the average spike. A 5 mm grid extended outside of the head was used as the initialisation points for the model. A non-linear search was then performed, starting from the optimal initial location.

## 3 Results

We recorded OPM data from four patients with mesial temporal lobe epilepsy while they performed a range of cognitive tasks in a magnetically shielded room, to assess whether interictal activity could be detected and localised with this novel neuroimaging method. The interictal results for all patients are shown in Figure 1. In total, 2, 9, 4 and 2 IEDs were detected for patients 1-4 respectively. The time series (all channels) and spatial topography (channels radial to the scalp) of the average of all interictal spikes are shown for each patient. The spatial topography is shown for the peak of the spike (t = 0 in the time series plot). The results of three different source localisation methods (an LCMV beamformer, a dipole fit to the peak of the spike and dipole fits at 2 ms intervals along the rising edge of the spike) are also shown, and exhibit reasonable agreement. Comparison of the source localisation with prior MRI and EEG recordings is given in Table 2. The interictal localisations for patients 1-3 agree well with previous clinical assessment, while the localisation for patient 4 is not concordant, with the MRI abnormality in the temporal lobe, while the OP-MEG localises to the frontal lobe. This may be due to the small number of IEDs observed.

**Table 2.** OP-MEG source localisation results, compared with prior clinical MRI and EEG.

| Participant No. | Findings from clinical MRI | Findings from clinical EEG | Interictal localisation | Ictal localisation |
| --- | --- | --- | --- | --- |
| 1 | Left Hippocampal sclerosis | No interictal; right inferior temporal ictal activity | Left temporal lobe | n/a |
| 2 | Left temporal lobe grey matter heterotopia | Left temporal interictal; left neocortical ictal | Left temporal lobe, 17.7 mm from lesion boundary | Left temporal lobe, 15.7 mm from lesion boundary |
| 3 | Normal | Right temporal | Right temporal lobe | Inferior frontal gyrus (R>L) |
| 4 | Left hippocampal sclerosis and left temporal focal cortical dysplasia | Left>Right temporal interictal; Right temporal ictal | Left frontal pole | n/a |

An ictal recording from patient 2 is shown in Figure 2. During this focal impaired awareness seizure, there is an observed increase in oscillatory activity within the 13-30 Hz range. There was very little participant movement in this time, moving only a maximum of 2.5 mm in the 2 s – 6 s period. An LCMV beamformer was used to localise this 13 Hz – 30 Hz activity, comparing power in the 13 Hz – 30 Hz frequency band in the ictal time window (2 s to 5 s in Figure 2) with power in the time window that followed (6 s to 9 s). As the ictal period was observed during a language task, to avoid biasing the results towards language areas of the brain, the covariance matrix was estimated from the participant’s resting recording. Otherwise the beamformer was the same as for the interictal data. This activity localised to the left temporal lobe, consistent with the previous clinical assessment and MRI findings, and with the peak 27.3 mm from the peak of the interictal activity found with the LCMV beamformer. We also looked at seizure progression, by evaluating the beamformer over 500 ms time windows. Based on this analysis, the ictal activity appears to begin close (14 mm in 0.5 – 1 s window) to the MRI lesion boundary and then spread anteriorly (distance between lesion boundary and localisation peak is 26 mm in 2.5 – 3 s window).

**Figure 2.**
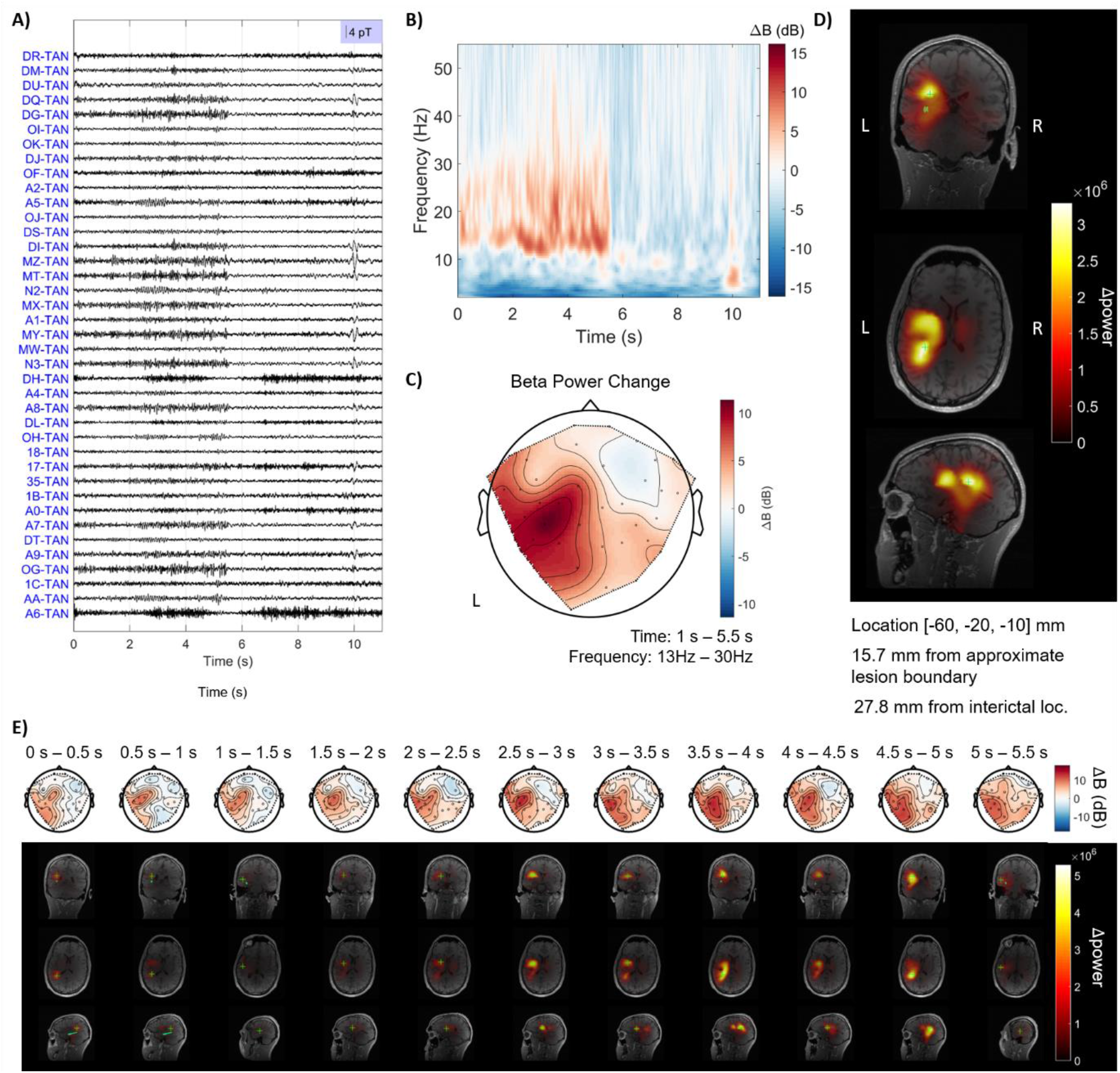
Ictal activity observed from patient 2. A) Time series. There is a visible increase in oscillatory activity between 2 – 6 s. There may also be a spike, as is often seen at the end of a seizure, at 10 s. B) Time-frequency spectrum of this ictal period of interest, averaged over channels and baseline corrected by all the average time-frequency spectrum for the rest of the recording. This oscillatory activity appears to be in the beta band (13 – 30 Hz). C) Topography (only on the OPM channels radial to the scalp) of the change in average beta power between 1 – 5.5 s during the ictal window and the remainder of the recording. It is largest over the left hemisphere. D) LCMV beamformer localisation of this beta power increase. The maximum power increase lies in the left temporal lobe and is marked by a green cross. The MRI abnormality is marked in green. E) LCMV beamformer for different time periods during the event. As in panel D, the MRI abnormality is shown in green.

An ictal recording for patient 3 is shown in Figure 3. This seizure was induced by playing music to the patient, while the clinician was in the room. The clinician pressed a button to mark the beginning of the clinical seizure in the OPM recording when the patient indicated by raising an arm that they were experiencing an aura. There was considerably more movement in this ictal event, with the patient exhibiting lip smacking (facial automatism) and the urge to turn their head to the left. As a result, large, low frequency signals are observed on the OPM channels, but it is difficult to disentangle the movement interference from the ictal signals of interest. Therefore, in Figure 3 we have focussed on observed high frequency increases, as these are unlikely to be induced by movement. To localise the activity, we used an LCMV beamformer to look at the power increase in the 60 Hz – 130 Hz range in the 11 s – 13 s time window by comparison with the 0 s – 2 s time window. Unlike the previous LCMV beamformer analyses, the source space was a 5 mm grid within the participant’s scalp surface (rather than the inner skull), as there was some concern that the high frequency signals could localise to the facial muscles. As seen in Figure 3, the power increase is bilateral, with a higher increase in the right hemisphere than left. The peak increase in power in the right hemisphere lies in the inferior frontal gyrus, while the peak of the left cluster lies in the orbitofrontal cortex. Both cluster peaks are adjacent to the respective superior temporal pole and superior temporal gyrus.

**Figure 3.**
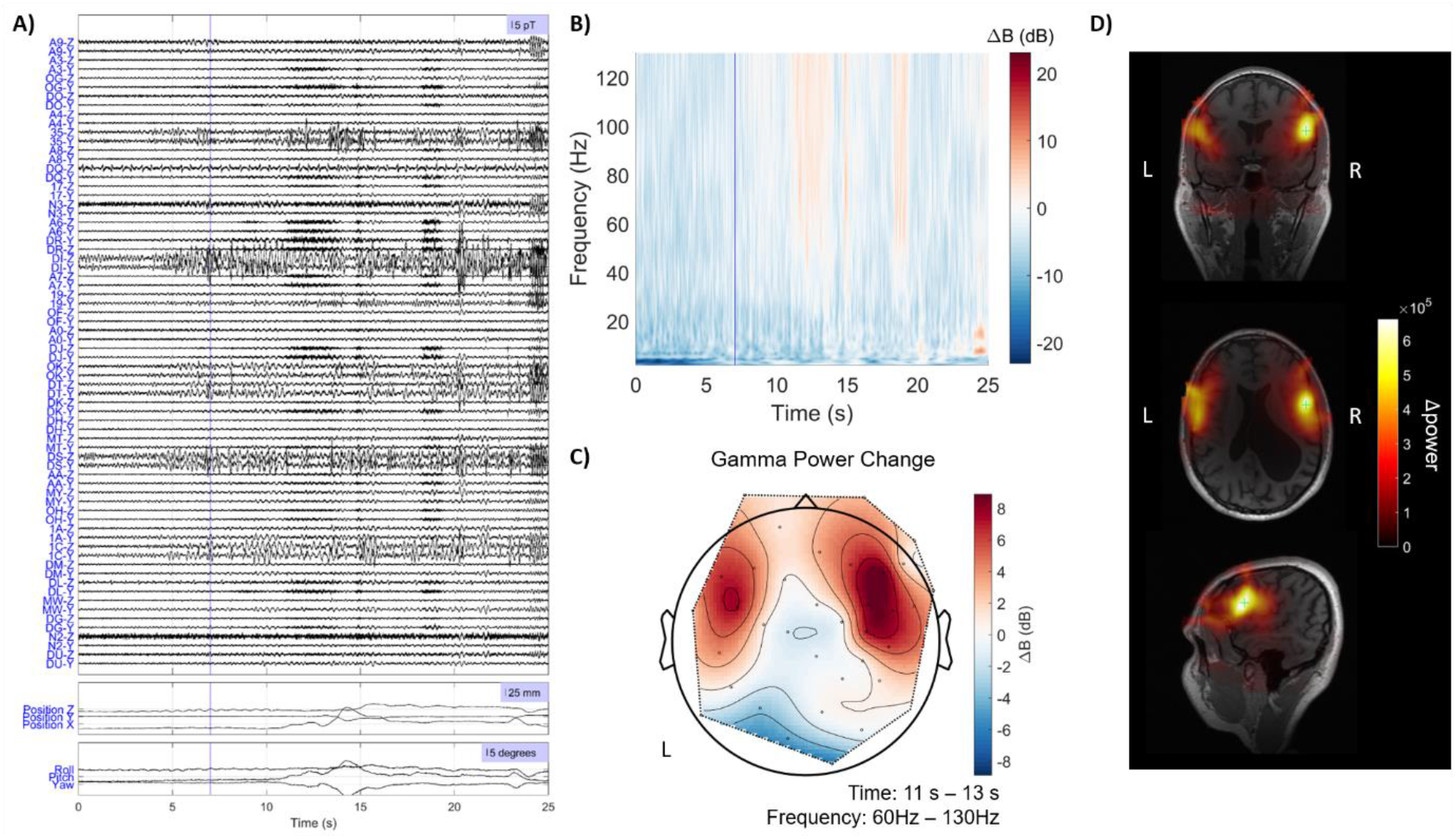
Ictal activity observed from patient 3. Patient reported commencement of aura at 7.02 s. A) Time series. Vertical blue line marks the start of the patient reported aura. There is a visible increase in variance in the sensor recordings at approximately 4 s. This may however be due to a combination of movement related field changes and the high-pass filter used in preprocessing the data, as there is an increase in movement at a similar time. B) Time-frequency spectrum of this time period, baseline corrected by the average of the data from this recording outside of this time period. C) Topography of the change in power in the 60 Hz – 130 Hz frequency range, between 11 s and 13 s, by comparison with all the data outside of this ictal window of interest. D) LCMV beamformer localisation. Increase in power in 60 Hz to 130 Hz frequency range between 11 s – 13 s by comparison with 0 s – 2 s. The green cross shows the peak power location.

## 4 Discussion

We have presented a small case series of mTLE recordings with OP-MEG. We have shown that mTLE can be visually identified from OP-MEG and localised to deep brain areas. These results support the clinical utility of OP-MEG. It is notable that no interictal discharges were observed in half of the patients. This is largely expected in an adult population and after OP-MEG recording, the patients’ previous clinical EEGs were examined. In two patients, these showed little interictal activity despite being telemetry recordings taken over days, unlike the short 30 minute to an hour OP-MEG recordings, suggesting that the absence of IEDs in the OP-MEG was due to natural heterogeneity between patients. However, as EEG was not recorded concurrently with OP-MEG and cryogenic MEG has not been recorded with these patients, this cannot be concluded with certainty.

We also present ictal OP-MEG for a subset of 2 of the patients. In one patient, ictal OP-MEG was recorded during a focal impaired awareness seizure. This shows clear activation localising to the expected region. The analysis is, however, slightly complicated by the fact that this ictal event was recorded during a language mapping task. In order to perform the source localisation, the covariance matrix of the data must be estimated. This requires at least approximately 45 s of data with similar noise properties to the event of interest. If the covariance matrix were estimated from the language data, the resulting localisation could be biased towards areas of the brain responsible for language generation. We therefore estimated the covariance matrix from the resting state recording and applied this to the ictal source localisation. This is a practical if imperfect solution, as the delay between the data from which the covariance matrix was estimated and the ictal event of interest may limit the noise reduction achieved by the beamformer.

We also recorded a seizure from a patient which was induced by listening to music. Here we have examined observed increases in high frequency activity during this seizure. These localised close to the superior temporal gyri, which is often associated with musicogenic seizures^16^. We cannot absolutely rule out that muscle artefacts could be the cause of these high frequency responses, but there was a high degree of participant movement during this recording and so we intentionally considered only high frequency (> 20 Hz) activity to minimise the impact of movement artefacts. Changes in low frequency signals would have been interesting to consider but could not be confidently disentangled from movement artefacts. Moving forward, this is likely to be less of a concern as there have recently been a number of developments in post-hoc software based interference correction for OP-MEG^13,17^. There were too few OPM channels in this recording to take advantage of these methods here however.

Future changes to OP-MEG recording setup could increase the rates of observed IEDs by undertaking some of the steps likely to increase epileptiform activity that would frequently be performed for a clinical EEG. These include activation procedures, reducing medication or sleep deprivation.

Additionally, recording time could be increased: these recordings were relatively short, ranging from 30 minutes to an hour. Foreseeably, OP-MEG could be used for considerably longer recordings, especially as personalised scanner-casts can be removed and replaced with minimal disruption to the recordings and sensor coregistration. This could improve success rates for patients and make the technology more widely applicable.

## 5 Conclusions

We have shown mesial temporal lobe epilepsies visually identified from OP-MEG. We show an example of how source localisation techniques can be used to increase detection of epileptogenic spikes and show localisation of both ictal and interictal events to the temporal lobe and hippocampal regions. This localisation is consistent with previous clinical assessment, demonstrating the potential of OP-MEG for presurgical planning for patients with mTLE.

## Data Availability

Anonymised OPM data will be made available upon request.

## References

1. Kuzmanovski I, Cvetkovska E, Babunovska M, et al. Seizure outcome following medical treatment of mesial temporal lobe epilepsy: Clinical phenotypes and prognostic factors. Clin Neurol Neurosurg. 2016;144:91–95. doi:10.1016/j.clineuro.2016.03.018

2. Wiebe S, Blume WT, Girvin JP, Eliasziw M. A Randomized, Controlled Trial of Surgery for Temporal-Lobe Epilepsy. N Engl J Med. 2001;345(5):311–318. doi:10.1056/NEJM200108023450501

3. Engel J, McDermott MP, Wiebe S, et al. Early Surgical Therapy for Drug-Resistant Temporal Lobe Epilepsy: A Randomized Trial. JAMA. 2012;307(9):922–930. doi:10.1001/jama.2012.220

4. Meckes-Ferber S, Roten A, Kilpatrick C, O’Brien TJ. EEG dipole source localisation of interictal spikes acquired during routine clinical video-EEG monitoring. Clin Neurophysiol. 2004;115(12):2738–2743. doi:10.1016/j.clinph.2004.06.023

5. Rampp S, Stefan H, Wu X, et al. Magnetoencephalography for epileptic focus localization in a series of 1000 cases. Brain. Published online 2019:3059–3071. doi:10.1093/brain/awz281

6. Schofield H, Boto E, Shah V, et al. Quantum enabled functional neuroimaging: the why and how of magnetoencephalography using optically pumped magnetometers. Contemp Phys. 2022;63(3):161–179. doi:10.1080/00107514.2023.2182950

7. Feys O, Corvilain P, Aeby A, et al. On-Scalp Optically Pumped Magnetometers versus Cryogenic Magnetoencephalography for Diagnostic Evaluation of Epilepsy in School-aged Children. Radiology. 2022;304(2):429–434. doi:10.1148/radiol.212453

8. Ruzich E, Crespo-García M, Dalal SS, Schneiderman JF. Characterizing hippocampal dynamics with MEG: A systematic review and evidence-based guidelines. Hum Brain Mapp. 2019;40(4):1353–1375. doi:10.1002/hbm.24445

9. Feys O, Ferez M, Corvilain P, et al. On-Scalp Magnetoencephalography Based On Optically Pumped Magnetometers Can Detect Mesial Temporal Lobe Epileptiform Discharges. Ann Neurol. 2024;95(3):620–622. doi:10.1002/ana.26844

10. Meyer SS, Bonaiuto J, Lim M, et al. Flexible head-casts for high spatial precision MEG. J Neurosci Methods. 2017;276:38–45. doi:10.1016/j.jneumeth.2016.11.009

11. Altarev I, Fierlinger P, Lins T, et al. Minimizing magnetic fields for precision experiments. J Appl Phys. 2015;117(23):233903. doi:10.1063/1.4922671

12. Litvak V, Mattout J, Kiebel S, et al. EEG and MEG data analysis in SPM8. Comput Intell Neurosci. 2011;2011:852961. doi:10.1155/2011/852961

13. Tierney TM, Seedat Z, St Pier K, Mellor S, Barnes GR. Adaptive multipole models of optically pumped magnetometer data. Hum Brain Mapp. 2024;45(4):e26596. doi:10.1002/hbm.26596

14. Tierney TM, Alexander N, Mellor S, et al. Modelling optically pumped magnetometer interference in MEG as a spatially homogeneous magnetic field. NeuroImage. 2021;244:118484. doi:10.1016/j.neuroimage.2021.118484

15. Nolte G. The magnetic lead field theorem in the quasi-static approximation and its use for magnetoencephalography forward calculation in realistic volume conductors. Phys Med Biol. 2003;48(22):3637. doi:10.1088/0031-9155/48/22/002

16. Stern J. Chapter 26 - Musicogenic epilepsy. In: Aminoff MJ, Boller F, Swaab DF, eds. Handbook of Clinical Neurology. Vol 129. The Human Auditory System. Elsevier; 2015:469–477. doi:10.1016/B978-0-444-62630-1.00026-3

17. Holmes N, Bowtell R, Brookes MJ, Taulu S. An Iterative Implementation of the Signal Space Separation Method for Magnetoencephalography Systems with Low Channel Counts. Sensors. 2023;23(14):6537. doi:10.3390/s23146537

