## Supplementary Table 1 for "Detection of mesial temporal lobe epilepsy with OP-MEG"

| Participant No. | Recording session | Paradigm | Recording Length (min) | Number of OPMs rejected | ICA components removed | IEDs detected |
|---|---|---|---|---|---|---|
| 1 | 1 | Rest (reading) | 10 | 0 | 7 | 0 |
| | | Rest (reading) | 10 | | 5 | 1 |
| | | Rest (without reading) | 10 | | 5 | 1 |
| | 2 | Memory | 21 | 0 | 3 | 0 |
| | | Verb Generation | 15 | | 3 | 0 |
| 2 | 1 | Rest | 10 | 1 | 3 | 1 |
| | | Rest | 10 | | 2 | 0 |
| | | Verb Generation | 14 | | 2 | 8 |
| 3 | 1 | Memory | 24 | 4 | 2 | 0 |
| | | Music | 11 | | 3 | 4 |
| 4 | 1 | Rest | 17 | 3 (2 Gen-2, 1 Gen-3) | 1 | 0 |
| | | Reward Processing | 22 | | 2 | 2 |

*Supplementary Table 1. Paradigm and analysis details for individual patients. ICA components were removed only for data inspection and spike detection, not for source localisation or any of the plots shown in the main manuscript.*
